# Residential bird biodiversity and frailty deficit accumulation: a longitudinal cohort study in New York City

**DOI:** 10.64898/2026.08.18.26360707

**Authors:** Pablo Knobel, Valentina Alaasam, Helena Krasnov, Itai Kloog, Vishal Midya, Alex Federman, Fred Ko, Maayan Yitshak Sade

**Affiliations:** Department of Environmental Medicine, Icahn School of Medicine at Mount Sinai, New York, New York, USA; Department of Biology, New York University, New York, New York, USA; Division of General Internal Medicine, Icahn School of Medicine, New York, NY, USA; Division of Geriatric Medicine, Larner College of Medicine, University of Vermont, Burlington, Vermont, USA

## Abstract

Urban nature is increasingly recognized as a determinant of healthy aging. However, research has largely focused on the quantity of greenness rather than biodiversity. Evidence supports an association between biodiversity and mental health, but physical aging evidence is very limited. We examined the longitudinal association between residential bird biodiversity and frailty severity using electronic health records. We conducted a retrospective cohort study of 20,388 adults aged 65 years and older receiving primary care in the Mount Sinai Health System in New York City, contributing 123,103 patient-years of follow-up (2011–2023). Residential bird biodiversity was derived from eBird citizen-science data as a modeled, bias-corrected latent Shannon diversity surface at the census-tract level yearly. Frailty severity was measured annually as the deficit count on the 31-item Veterans Affairs Frailty Index (VA-FI). We estimated associations using a negative binomial generalized additive model adjusted for age, sex, race and ethnicity, insurance, tract-level poverty, and non-Hispanic Black proportion, reporting results as the percent change in expected deficit count. We tested effect modification by age group (65–74, 75–84, ≥85 years). Each interquartile range increase in residential bird Shannon diversity was associated with a 1.4% lower expected VA-FI deficit count (95% CI −2.1% to −0.8%). The association was strongest among adults aged 65–74 years (−3.0%, 95% CI −3.9% to −2.1%), attenuated among those aged 75–84 years (−0.8%, 95% CI −1.9% to 0.3%), and no longer evident among those aged 85 and older (+1.6%, 95% CI −0.0% to 3.3%). Greater residential bird biodiversity (reflecting both species richness and evenness) was associated with lower frailty severity, with the largest association in early old age. As a bioindicator of underlying environmental quality shaped by modifiable urban design, bird diversity may point to a avenue for supporting healthy aging in dense cities.

## 1. Introduction

The global population is aging at an unparalleled pace. The ratio of individuals aged 65 and older to those under 65 is projected to rise from 1:11 to 1:6 by 2050, representing a growth from 703 million to 1.5 billion older adults worldwide^1^. The majority of this growth will occur in urban areas, where environmental conditions are shaped by human-made infrastructure and policy decisions, making cities both a key determinant of, and a critical venue for, promoting healthy aging^2^. Among the most consequential manifestations of aging is frailty, defined by cumulative decline across multiple physiological systems resulting in heightened vulnerability to adverse health events^3^. Frailty is associated with increased risk of disability, hospitalization, and mortality, and imposes a substantial and growing burden over healthcare systems globally^4^.

Urban nature has emerged as a promising environmental determinant of healthy aging. A growing body of evidence links residential greenness to improved aging-related outcomes, including cognitive function, physical capability, and perceived well-being^5^. There is some evidence for the associations between urban greenness and frailty^6-8^, all from studies in East Asia. However, most of this research focuses on the geographic distribution of urban greenness, omitting variation in vegetation diversity, ecological function, connectivity, and the quality of human-nature interactions, factors increasingly recognized as central to health-promoting mechanisms^9^. This oversimplification limits our ability to identify what specific features of urban nature confer benefit^10^. A second limitation is practical: greenness-focused approaches implicitly assume that expanding green area is the primary lever for improving health, a strategy poorly suited to densely built cities where land is in high demand.

Evidence suggests that bird biodiversity provides diverse benefits to human health, including direct psychological and physiological benefits^11^. Exposure to birdsong has been shown to reduce anxiety and alleviate attentional fatigue^12,13^, and exposure to greater bird species richness has been associated with improved psychological well-being among urban green space users^14^. Some observational epidemiological studies support these associations: Chen et al. (2023)^15^ found that higher bird species richness was associated with longer life expectancy and lower cause-specific mortality across 2,751 US counties after adjustment for socioeconomic and healthcare covariates. Buxton et al. (2024)^16^ reported that residing in a Canadian postal code with higher bird diversity was associated with a 6.64% increase in self-reported good mental health. Bird biodiversity is also a modifiable exposure: urban nature initiatives including habitat restoration, reduced light and noise pollution, and native planting can meaningfully increase local bird diversity^17^. Indeed, cities can support high avian diversity, in some cases exceeding that of surrounding peri-urban or rural landscapes, providing an especially promising context for this exposure^18^.

A key barrier to the use of bird biodiversity metrics in epidemiologic studies is that available biodiversity metrics are too coarse to reflect the variation in biodiversity that can occur within small areas^19^. Some studies have used citizen science platforms such as eBird to generate finer-resolution exposure estimates, for example, aggregating to postal code or census tract rather than ZIP code or county, but these approaches have not addressed the inherent limitations of citizen science data itself, including uneven and self-selected observer effort, variable observer skill, and irregular spatial and temporal sampling coverage, which can bias biodiversity estimates independent of true ecological variation^20^. Bird biodiversity functions as a direct experiential exposure and as a bioindicator of other environmental determinants of aging, like air, light, or noise pollution that function as direct exposures^21^. At coarse spatial units such as ZIP codes or counties, a biodiversity measure largely reflects that general environmental quality rather than the biodiversity people encounter around their homes, obscuring any direct contribution of species diversity itself.

In this study, we developed a bias-corrected bird biodiversity model for New York City census tracts using eBird citizen science data, and linked it to electronic health records from 20,388 older adults receiving primary care in the Mount Sinai Health System. Using residential addresses and 123,103 patient-years of follow-up between 2011 and 2023, we estimated the association between residential bird biodiversity and frailty severity, measured as the count of accumulated health deficits on the Veterans Affairs Frailty Index, and tested whether that association differs across the life course. To our knowledge, this is the first study to examine avian biodiversity in relation to a clinical measure of physical aging, and the first to do so with a time-varying exposure at the neighborhood scale.

## 1. Methods

### 2.1 Study population

Our retrospective cohort study included 20,388 New York City residents over 65 years (123,103 patient-years of follow-up) within the Mount Sinai Health System (MSHS) from 2011 to 2023. Electronic health record (EHR) data were obtained from the Mount Sinai Data Warehouse. For cohort construction, we identified each participant’s first documented encounter within MSHS as their baseline year and subsequently followed them annually until death, censoring at 2023, or if they had two consecutive years in which no healthcare utilization was recorded, with the censoring year defined as the last year in which any service use was documented. Demographic data, including age, sex, race/ethnicity, and insurance information, were obtained from EHRs. We excluded participants without a valid residential address (0.5%) or who lacked valid covariate data (5.4%). This study was approved by the Institutional Review Board of Mount Sinai (STUDY 20–01498), and a waiver of informed consent was granted.

### 2.2 Exposure Assessment: Latent Bird Biodiversity Surface

Bird observations were drawn from eBird, a citizen-science platform maintained by the Cornell Lab of Ornithology^22^. eBird users, predominantly amateur birdwatchers, submit checklists through the eBird app or website each time they go birding. Checklists are a record of every species seen or heard during a single outing, along with the date, location, duration, and distance covered. A single location can accumulate many checklists over time, submitted by different observers on different visits, rather than reflecting one fixed monitoring station. Although the United States is among the highest-quality eBird environments^23^, the data carry well-documented spatial biases: observations cluster near roads, trails, and wealthier, predominantly white neighborhoods^24^(**Figure 1B**). Therefore, assigning raw counts to residential addresses yields uneven, biased exposure estimates. Therefore, we modeled a latent biodiversity surface that recovers the underlying ecological signal while correcting for sampling effort and coverage.

**Figure 1.**
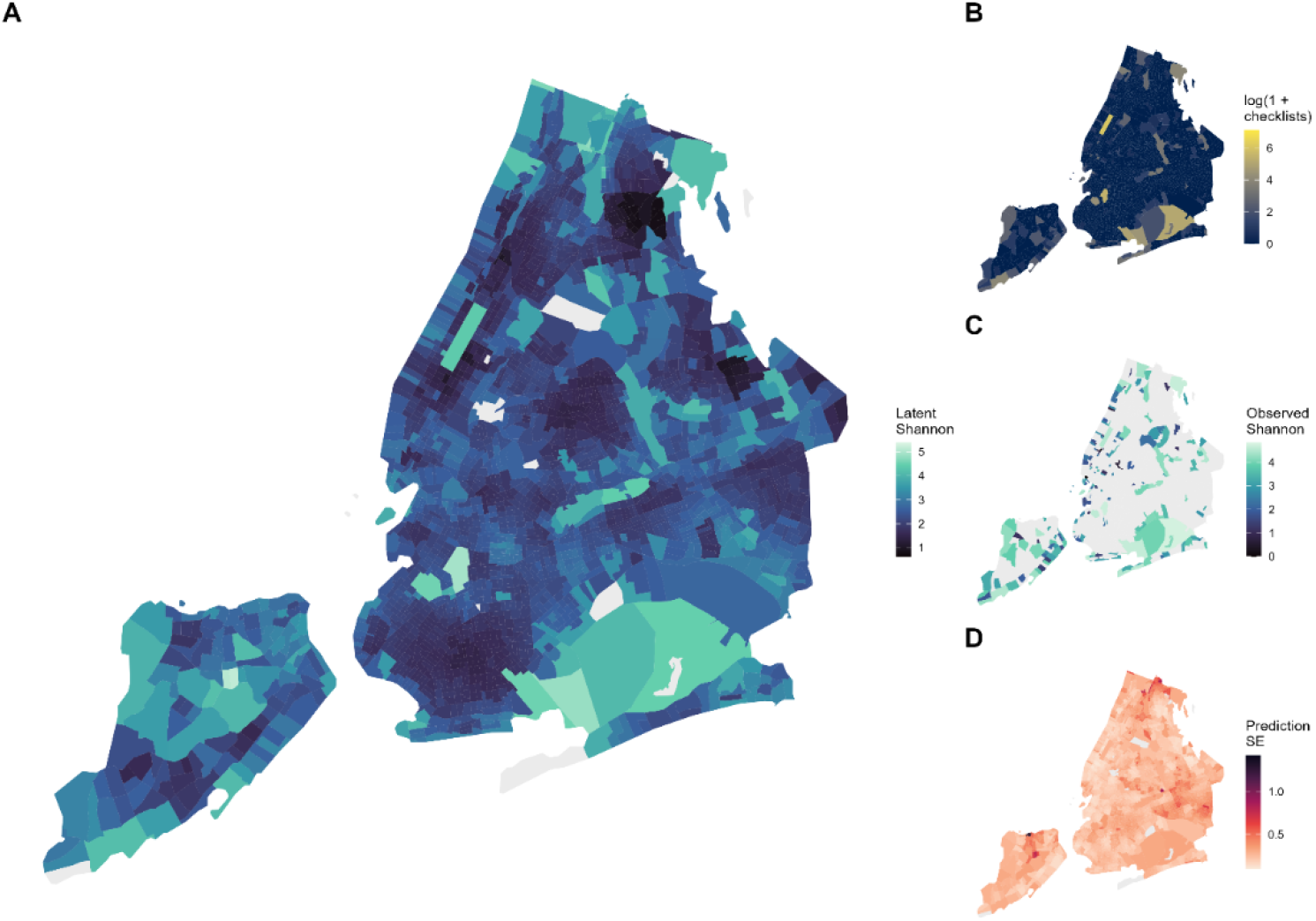
**Panel A**. GAM-estimated latent bird Shannon index across NYC census tracts. **Panel B**. eBird checklist density (log scale) per census tract. Darker tracts received more checklists; grey tracts had no observations. **Panel C**. Raw checklist-derived Shannon diversity index per census tract. Grey tracts had insufficient eBird coverage to compute diversity. **Panel D**. Model prediction uncertainty (standard error). Lighter shading indicates greater confidence in the latent diversity estimate. All panels are for 2017 as an example.

We used New York City census tracts to construct the biodiversity exposure surface. New York City comprises 34,905 possible census tract-years over the study period (2,327 tracts × 2010-2024 years). We restricted analysis to complete checklists (i.e. eBird on which the observer recorded every bird species detected, rather than only notable or target species) to compute a diversity estimate directly. Only 2,528 tract-years (7.2%) had adequate checklists for inclusion (Figure 1C). Within each included tract-year, we calculated species-level encounter rates (i.e. the proportion of complete checklists on which each species was detected) and used these to compute Shannon diversity for that tract-year. We partitioned these into a training set of 2022 tract-years (80%) and a held-out test set of 506 tract-years (20%). We additionally excluded tracts averaging fewer than 100 inhabitants (n=4.4%), which are predominantly non-residential urban parks rather than places of residence; these were retained only as spatial-lag sources for predictors.

To generate a spatially and temporally resolved latent biodiversity surface, we fitted a Generalized Additive Model (GAM) with a Gamma family and log link to accommodate the right-skewed, positive-valued distribution of Shannon diversity indices. The model included a two-dimensional spatial smooth over census-tract centroids to capture spatial gradients in bird biodiversity, a temporal smooth over years to model trends, including changes in eBird participation over time, and a ZIP code random effect to absorb residual observer clustering. All additional predictors were entered with penalized regression splines and subject to automatic smoothness selection. Predictor variables were selected to represent the key ecological and environmental determinants of urban bird diversity and were drawn from nationally available data sources to ensure reproducibility, scalability, and temporal extension. Land cover was characterized using the National Land Cover Database^25^. Vegetation greenness was measured using the Normalized Difference Vegetation Index (NDVI)^26^. Urban-rural character was captured using Rural-Urban Commuting Area (RUCA) codes^27^. Food environment quality was characterized using the Modified Retail Food Environment Index (mRFEI)^28^. Sociodemographic context was included via American Community Survey (ACS) variables^29^, and ambient air quality was represented using modeled PM_2.5_ components data. We imputed missing predictor values. Spatial lag predictors were computed using first-and second-order contiguity to capture spillover effects from neighboring tracts.

Predictive accuracy was assessed in a held-out test set of 506 census tract-years using two complementary metrics. The coefficient of determination (R^2^ = 0.558) indicates the model explained approximately 56% of the variance in observed Shannon diversity values in unseen data. Spearman’s rank correlation (ρ = 0.745) indicates the model also preserved the relative ordering of tracts from lowest to highest diversity (**Figure 1A**). This performance is broadly comparable to a national-scale model of bird species richness across the contiguous United States, which used random forest models at coarser (0.5–5 km) resolutions and reported percentage variance explained ranging from 27% to 60% (median 54%) across spatial resolutions^30^. Each participant was assigned the predicted Shannon diversity value corresponding to their residential census tract and calendar year of follow-up, yielding a time-varying exposure. For all analyses, exposure was expressed as the interquartile range (IQR) of the tract-level Shannon diversity distribution.

### 2.3 Outcome: Veterans Affairs Frailty Index

Frailty status was operationalized using the Veterans Affairs Frailty Index (VA-FI)^4^, a 31-item cumulative deficit index constructed from diagnostic (ICD-10) and procedure (CPT) codes in EHRs. The VA-FI was developed using the cumulative deficit method described by Searle et al.^3^, in which variables are included if they meet four criteria: association with health status, increasing prevalence with age, absence of saturation before age 65, and coverage of diverse physiological systems. Each deficit was assessed annually using a three-year lookback window (the index year and the two preceding years); deficits with no qualifying codes during that window were coded as absent. While the VA-FI was developed and validated in the Veterans Health Administration population, its construction relies solely on standard ICD-10 diagnostic and CPT procedure codes present in our EHR system. While the VA-FI score is conventionally expressed as the proportion of deficits present out of the total number of items, we used the deficit count^31^.

#### Statistical Analysis

We estimated the association between annual residential bird biodiversity, using the latent Shannon diversity index, and frailty deficit count. We used a negative binomial generalized additive model (GAM) including a penalized spline of calendar year, and a patient-level random intercept to account for repeated measures. We adjusted the model for age, sex, race and ethnicity, insurance, tract-level poverty, and, to capture neighborhood racial composition independent of individual patient characteristics, tract-level percentage of non-Hispanic Black residents. We exponentiated each coefficient to a count ratio and report it as the percent change in expected deficit count with 95% confidence intervals.

In a secondary analysis, we tested effect modification by age by adding an interaction between IQR-scaled Shannon diversity and age group (65–74 / 75–84 / 85+ years) to the primary model. Each age stratum’s association was obtained as a linear combination of the Shannon main-effect coefficient and the corresponding age-group interaction term (for the reference stratum, the main effect alone), with standard errors computed from the model’s variance–covariance matrix.

We conducted two sensitivity analyses to evaluate the robustness of the primary model. SA1 refit the primary model with inverse-variance weighting by the uncertainty of the tract-level biodiversity predictions, so that participants in more precisely estimated tracts contributed more heavily (**Figure 1D**), to evaluate whether this weighting scheme changed the association observed in the unweighted primary model. SA2 refit the primary model using a quasi-Poisson specification in place of the negative binomial, to confirm that the overdispersion-handling approach did not materially affect the estimated association.

## 2. Results

The analytical sample comprised 20,388 patients aged 65 years and older, contributing 123,103 patient-years of follow-up between 2011 and 2023. At cohort entry, the mean age was 71.3 years (SD 7.2), and 64.7% of patients were female. In terms of race and ethnicity, 45.1% were White, 14.5% were Black, and 3.9% were Asian. Insurance coverage was predominantly Medicare (73.3%), followed by Commercial (16.8%), Other (7.2%), and Medicaid (2.7%). The mean percentage of census tract residents living in poverty was 16.2% (SD 11.9), and the mean percentage identifying as non-Hispanic Black was 15.7% (SD 20.7). The mean IQR-scaled Shannon diversity index at entry was 3.04 (SD 0.83). The mean VAFI deficit count at entry was 1.74 (SD 2.56) (**Table 1**).

**Table 1.**
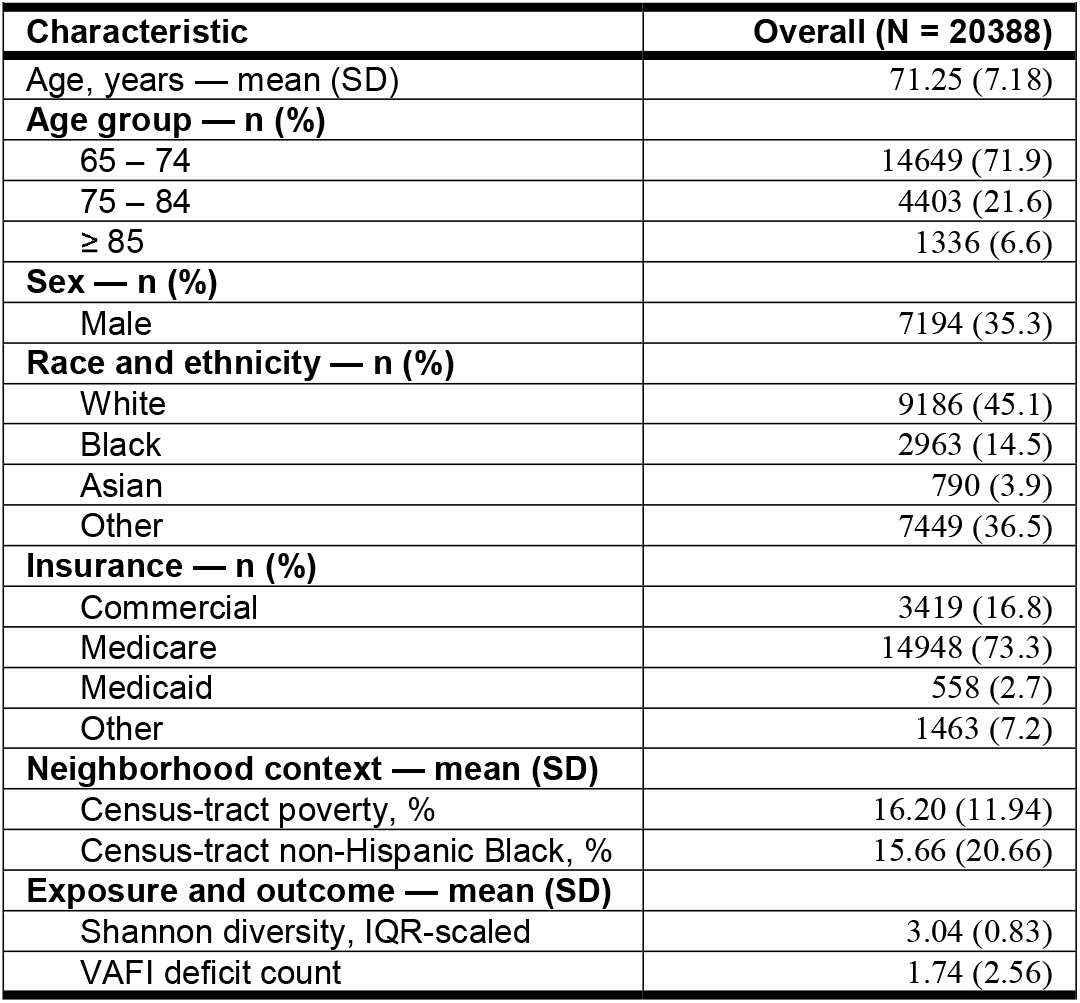
Characteristics at cohort entry.

| **Table 1. Characteristics at cohort entry** |  |
| --- | --- |
| **Characteristic** | **Overall (N = 20388)** |
| Age, years — mean (SD) | 71.25 (7.18) |
| **Age group — n (%)** |  |
| 65 – 74 | 14649 (71.9) |
| 75 – 84 | 4403 (21.6) |
| ≥ 85 | 1336 (6.6) |
| **Sex — n (%)** |  |
| Male | 7194 (35.3) |
| **Race and ethnicity — n (%)** |  |
| White | 9186 (45.1) |
| Black | 2963 (14.5) |
| Asian | 790 (3.9) |
| Other | 7449 (36.5) |
| **Insurance — n (%)** |  |
| Commercial | 3419 (16.8) |
| Medicare | 14948 (73.3) |
| Medicaid | 558 (2.7) |
| Other | 1463 (7.2) |
| **Neighborhood context — mean (SD)** |  |
| Census-tract poverty, % | 16.20 (11.94) |
| Census-tract non-Hispanic Black, % | 15.66 (20.66) |
| **Exposure and outcome — mean (SD)** |  |
| Shannon diversity, IQR-scaled | 3.04 (0.83) |
| VAFI deficit count | 1.74 (2.56) |

In the primary negative binomial GAM, each one-IQR increase in residential bird Shannon diversity was associated with a 1.4% lower expected VAFI deficit count across follow-up (95% CI −2.1% to −0.8%). The association was similar when inverse-variance weighting was applied (SA1: −1.4%, 95% CI −1.5% to −1.2%) and under a quasi-Poisson specification (SA2: −1.5%, 95% CI −2.1% to −0.8%) (**Table 2**).

**Table 2.**
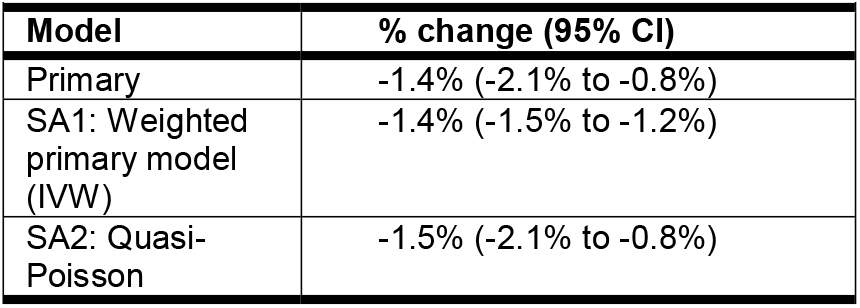
Primary and sensitivity analysis results.

| **Table 2: Primary and sensitivity analysis results** |  |
| --- | --- |
| **Model** | **% change (95% CI)** |
| Primary | -1.4% (-2.1% to -0.8%) |
| SA1: Weighted primary model (IVW) | -1.4% (-1.5% to -1.2%) |
| SA2: Quasi-Poisson | -1.5% (-2.1% to -0.8%) |

The association between bird biodiversity and frailty severity varied by age group (**Table 3**). It was strongest among patients aged 65 to 74 years (−3.0%, 95% CI −3.9% to −2.1%) and diminished among those 75 to 84 (−0.8%, 95% CI −1.9% to 0.3%). Among patients 85 and older, the association was no longer evident (+1.6%, 95% CI −0.0% to 3.3%)

**Table 3.** Shannon diversity and VAFI deficit count by age group.

| Age group | % change (95% CI) |
| --- | --- |
| 65–74 | -3.0% (-3.9% to -2.1%) |
| 75–84 | -0.8% (-1.9% to 0.3%) |
| 85+ | 1.6% (-0.0% to 3.3%) |

## 3. Discussion

In this longitudinal study of 20,388 adults aged 65 and older receiving primary care in New York City, greater residential bird biodiversity was associated with lower frailty severity overall (−1.4%, 95% CI −2.1% to −0.8%). This association was strongest among adults aged 65 to 74 and progressively disappeared at older ages.

Our finding of a protective association between bird biodiversity and frailty severity is consistent with an emerging literature demonstrating beneficial associations between bird species diversity and health outcomes. Methorst et al. (2021)^32^ found that bird species richness was positively associated with life satisfaction across 26 European countries in data from over 26,000 respondents. Most closely analogous to our design, Buxton et al. (2024)^16^ linked eBird-derived bird Shannon diversity to self-rated mental health across 36 Canadian metropolitan areas using a longitudinal survey spanning 2007–2022; postal codes with bird diversity one standard deviation above the mean showed approximately 6.6% higher odds of reporting good mental health. While this is the first study to investigate the association between avian biodiversity and frailty specifically, a parallel literature already links the broader urban-nature exposure of residential greenness to frailty in older adults. Using the Chinese Longitudinal Healthy Longevity Survey, Zhu et al. (2020)^6^ found that older adults in the greenest residential quartile had 14% lower odds of frailty than those in the least green quartile. Extending this beyond vegetation quantity alone, He et al. (2022)^7^ showed in the same cohort that larger, more complex, and more connected greenspace structures were associated with lower frailty, particularly among women and the oldest-old. In Los Angeles, Chen et al. (2025)^33^ demonstrated that bird species richness partially mediated the association between greenspace percentage and prevalence of poor mental health across 294 census tracts, suggesting that avian diversity may be an active pathway through which urban greenspace confers health benefits.

Experimental and quasi-experimental evidence on bird-specific stimuli reinforces the plausibility of the observed association. Stobbe et al. (2022)^34^ showcased, in a randomized online experiment with 295 participants, that exposure to birdsong soundscapes significantly reduced anxiety and paranoia. Similarly, Ratcliffe et al. (2013)^12^ found through semi-structured interviews that birdsong was the natural sound most commonly associated with perceived stress recovery and attention restoration, with restorative appraisals linked to the acoustic, aesthetic, and associative properties of bird sounds. Van Hedger et al. (2019)^35^ showed that brief exposure to natural soundscapes including animal sounds improved directed attention performance relative to urban soundscapes. These experimental findings on psychological and cognitive restoration suggest plausible mechanistic links between residential bird biodiversity and the functional deficits captured by the VAFI, several of which tap domains such as cognition, mood, and physical functioning that are sensitive to chronic stress and attentional fatigue.

The protective association was not uniform across the life course. It was strongest among adults aged 65 to 74 years (−3.0%, 95% CI −3.9% to −2.1%) and attenuated among those 75 to 84 (−0.8%, 95% CI −1.9% to 0.3%). Among adults 85 and older, the association was no longer present (+1.6%, 95% CI −0.0% to 3.3%). This aligns with a broader literature on urban nature and healthy aging: a recent systematic review of green space and older-adult health concluded that benefits are most pronounced among “younger seniors” aged 65 to 74, a pattern attributed to their greater mobility and reliance on local green space for recreation, physical activity, and social connection^36^. Among adults 75 and older, declining life-space mobility likely reduces direct contact with the local avian environment ^37^, while the accelerating, multimorbidity-driven deficit accumulation characteristic of advanced age may overwhelm the comparatively modest contribution of neighborhood biodiversity^38^. The 65-to-74 window may therefore mark a period in which older adults remain mobile enough to engage with neighborhood nature yet have begun to accrue the deficits on which such exposure can act.

Our work extends the literature in four ways. First, it examines a clinical measure of physical aging rather than self-rated mental health. Moreover, VAFI captures a broad spectrum of frailty-relevant deficits across physical, functional, and cognitive domains, enabling analysis of deficit accumulation as a continuous biological process rather than binary frailty status alone. Second, it employs a time-varying exposure linked to annual residential location rather than cross-sectional assignment. Third, our exposure is itself derived from a modeled, bias-corrected biodiversity surface built from eBird citizen science. Fourth, it does so in a large, sociodemographically heterogeneous EHR cohort in a dense U.S. city, with up to 13 years of follow-up, offering greater population representativeness than the convenience samples characteristic of experimental studies.

This study has several limitations. First, we measured bird biodiversity at the residential census-tract level, which may not reflect the outdoor spaces they use most or where they spend most of the time. However, birds sing most at dawn and dusk. These are times when people are most likely to be at home. Second, although we adjusted for a wide range of socioeconomic, demographic, and land-cover variables, we cannot rule out residual confounding from factors we did not measure, such as built-environment quality, social capital, and early-life environmental exposures. Even so, the variables we adjusted for account for the main confounders proposed in this literature, and the association remained consistent across sensitivity analyses that varied both the exposure window and the follow-up period. Finally, because the VAFI is derived from the EHR, its trajectories reflect both changes in health and the frequency with which patients engage with the health system. Differential healthcare use across tracts with different biodiversity cannot be fully ruled out as a source of bias. To limit this, we ended follow-up once patients stopped using the health system, retaining only periods of active care during which deficits could reliably be recorded.

## 4. Implications and conclusion

To our knowledge, this is the first longitudinal epidemiological study to examine the association between residential bird biodiversity and frailty in an urban population. Our findings support the hypothesis that greater residential bird diversity is associated with slower accumulation of functional deficits in older adults, independent of socioeconomic and demographic covariates, and that this association may be moderated by age, with the largest benefit observed among adults aged 65 to 74 years. Future work should seek to identify the behavioral and physiological mediators of this relationship, examine whether biodiversity-promoting interventions slow frailty progression in randomized controlled trials, and evaluate the distributional equity of biodiversity-health benefits across sociodemographic groups. This study contributes to a growing body of evidence suggesting that urban biodiversity represents a modifiable environmental determinant of health in older adults: one that may be amenable to intervention through urban greening, habitat restoration, and land-use policy.

## Data Availability

Exposure data in the present study are available upon reasonable request to the authors. Health data is availible upon reasonable request to the Mount Sinai Data Wharehouse.

## Acknowledgments

This study was supported by the Mount Sinai Transdisciplinary Center on Early Environmental Exposures (P30 ES023515), and through the computational and data resources and staff expertise provided by Scientific Computing and Data at the Icahn School of Medicine at Mount Sinai. VJA is supported by the NSF Postdoctoral Research Fellowships in Biology Program under Grant No. 2305367.

## References

1. United Nations DoEaSA, Population Division World Population Prospects 2022: Summary of Results. Vol. UN DESA/POP/2022/TR/NO. 3. 2022.

2. Wood G, Pykett J, Daw P, et al. The role of urban environments in promoting active and healthy aging: A systematic scoping review of citizen science approaches. Journal of Urban Health. 2022;99(3):427–456.

3. Searle SD, Mitnitski A, Gahbauer EA, Gill TM, Rockwood K. A standard procedure for creating a frailty index. BMC geriatrics. 2008;8(1):1–10.

4. Cheng D, DuMontier C, Yildirim C, et al. Updating and validating the US Veterans Affairs Frailty Index: transitioning from ICD-9 to ICD-10. The Journals of Gerontology: Series A. 2021;76(7):1318–1325.

5. de Keijzer C, Bauwelinck M, Dadvand P. Long-term exposure to residential greenspace and healthy ageing: A systematic review. Current environmental health reports. 2020;7(1):65–88.

6. Zhu A, Yan L, Wu C, Ji JS. Residential greenness and frailty among older adults: a longitudinal cohort in China. Journal of the American Medical Directors Association. 2020;21(6):759-765. e2.

7. He Q, Chang H-T, Wu C-d, Ji JS. Association between residential greenspace structures and frailty in a cohort of older Chinese adults. Communications medicine. 2022;2(1):43.

8. Yu R, Wang D, Leung J, Lau K, Kwok T, Woo J. Is neighborhood green space associated with less frailty? Evidence from the Mr. and Ms. Os (Hong Kong) study. Journal of the American Medical Directors Association. 2018;19(6):528–534.

9. Jarvis I, Gergel S, Koehoorn M, van den Bosch M. Greenspace access does not correspond to nature exposure: Measures of urban natural space with implications for health research. Landscape and Urban Planning. 2020;194:103686.

10. Marselle MR, Hartig T, Cox DT, et al. Pathways linking biodiversity to human health: A conceptual framework. Environment International. 2021;150:106420.

11. Gray A, Doyle S, Doyle C, Young JC, McMahon BJ. Birds and human health: Pathways for a positive relationship and improved integration. Ibis. 2024;166(3):761–779.

12. Ratcliffe E, Gatersleben B, Sowden PT. Bird sounds and their contributions to perceived attention restoration and stress recovery. Journal of Environmental Psychology. 2013;36:221–228. doi:10.1016/j.jenvp.2013.08.004

13. Stobbe E, Sundermann J, Ascone L, Kühn S. Birdsongs alleviate anxiety and paranoia in healthy participants. Scientific Reports. 2022;12(1):16414.

14. Dallimer M, Irvine KN, Skinner AM, et al. Biodiversity and the feel-good factor: understanding associations between self-reported human well-being and species richness. BioScience. 2012;62(1):47–55.

15. Chen Y, Zhao P, Xu Q, et al. Relating biodiversity with health disparities of human population: An ecological study across the United States. One Health. 2023;16:100548.

16. Buxton RT, Hudgins EJ, Lavigne E, et al. Mental health is positively associated with biodiversity in Canadian cities. Commun Earth Environ. 2024;5(1):310. doi:10.1038/s43247-024-01482-9

17. Ciach M, Fröhlich A. Habitat type, food resources, noise and light pollution explain the species composition, abundance and stability of a winter bird assemblage in an urban environment. Urban Ecosystems. 2017;20:547–559.

18. Dearborn DC, Kark S. Motivations for conserving urban biodiversity. Conservation biology. 2010;24(2):432–440.

19. Fraixedas S, Lindén A, Piha M, Cabeza M, Gregory R, Lehikoinen A. A state-of-the-art review on birds as indicators of biodiversity: Advances, challenges, and future directions. Ecological Indicators. 2020;118:106728.

20. Callaghan CT, Gawlik DE. Efficacy of eBird data as an aid in conservation planning and monitoring. Journal of Field Ornithology. 2015;86(4):298–304.

21. Pollack L, Ondrasek NR, Calisi R. Urban health and ecology: the promise of an avian biomonitoring tool. Curr Zool. Apr 2017;63(2):205–212. doi:10.1093/cz/zox011

22. Sullivan BL, Wood CL, Iliff MJ, Bonney RE, Fink D, Kelling S. eBird: a citizen-based bird observation network in the biological sciences. Biological Conservation. 2009;142:2282–2292.

23. La Sorte FA, Somveille M. Survey completeness of a global citizen □ science database of bird occurrence. Ecography. 2020;43(1):34–43.

24. Grade AM, Chan NW, Gajbhiye P, Perkins DJ, Warren PS. Evaluating the use of semi-structured crowdsourced data to quantify inequitable access to urban biodiversity: A case study with eBird. Plos one. 2022;17(11):e0277223.

25. Survey USG. Data from: Annual National Land Cover Database (NLCD) Collection 1 Products (ver. 1.2, June 2026). 2024. Sioux Falls, SD. doi:10.5066/p94uxnts

26. Rugel EJ, Henderson SB, Carpiano RM, Brauer M. Beyond the Normalized Difference Vegetation Index (NDVI): Developing a Natural Space Index for population-level health research. Environ Res. Nov 2017;159:474–483. doi:10.1016/j.envres.2017.08.033

27. U.S. Department of Agriculture ERS. Data from: 2020 Rural-Urban Commuting Area Codes. 2025. Washington, DC.

28. Division of Nutrition, Physical Activity,, and Obesity, National Center for Chronic Disease Prevention and Health Promotion, CDC,. Census Tract Level State Maps of the Modified Retail Food Environment Index (mRFEI). https://www.cdc.gov/obesity/downloads/census-tract-level-state-maps-mrfei_TAG508.pdf

29. U.S. Census Bureau. Explore Census Data. https://data.census.gov/

30. Carroll KA, Farwell LS, Pidgeon AM, et al. Mapping breeding bird species richness at management □ relevant resolutions across the United States. Ecological Applications. 2022;32(6):e2624.

31. Krasnov H, Hung W, Knobel P, et al. Urban Exposures, Frailty, and Mental Illness in World Trade Center Health Program Responders. medRxiv. 2026;

32. Methorst J, Bonn A, Marselle M, Böhning-Gaese K, Rehdanz K. Species richness is positively related to mental health – A study for Germany. Landscape and Urban Planning. 2021;211 doi:10.1016/j.landurbplan.2021.104084

33. Chen S, Wang H, Xu W. Bird richness as a mediator between greenspace and mental health relationships. Landscape and Urban Planning. 2025;259 doi:10.1016/j.landurbplan.2025.105360

34. Stobbe E, Sundermann J, Ascone L, Kuhn S. Birdsongs alleviate anxiety and paranoia in healthy participants. Sci Rep. Oct 13 2022;12(1):16414. doi:10.1038/s41598-022-20841-0

35. Van Hedger SC, Nusbaum HC, Clohisy L, Jaeggi SM, Buschkuehl M, Berman MG. Of cricket chirps and car horns: The effect of nature sounds on cognitive performance. Psychon Bull Rev. Apr 2019;26(2):522–530. doi:10.3758/s13423-018-1539-1

36. Wang M, Che Y, Tan X, Zhang N, Yu S, Yan P. Association between green space exposure and elderly health: a systematic review and meta-analysis. BMC Public Health. 2026;

37. Rantakokko M, Mänty M, Rantanen T. Mobility decline in old age. Exercise and sport sciences reviews. 2013;41(1):19–25.

38. Kaskirbayeva D, West R, Jaafari H, et al. Progression of frailty as measured by a cumulative deficit index: a systematic review. Ageing Research Reviews. 2023;84:101789.

